# Forecasting PPE Consumption during a Pandemic: The Case of Covid-19

**DOI:** 10.1101/2020.08.20.20178780

**Authors:** Kristian Lum, James Johndrow, April Cardone, Barry Fuchs, Cody E. Cotner, Olivia Jew, Ravi B. Parikh, Michael Draugelis, ThaiBinh Luong, Asaf Hanish, Gary E. Weissman, Christian Terwiesch, Kevin G. Volpp

## Abstract

Due to the global shortage of PPE caused by increasing number of COVID-19 patients in recent months, many hospitals have had difficulty procuring adequate PPE for the clinicians who care for these patients. Faced with a shortage, hospitals have had to implement new PPE conservation policies. In this paper, we describe a tool to help hospitals better project PPE needs under various conservation policies. Though this tool is built on top of projections of the number of hospitalized COVID-19 patients, it is agnostic as to which model—of which many are available—provides these projections. The tool combines COVID-19 patient census projections with information like staffing ratios and frequency of patient contact to provide projections of the number of items of key types of PPE needed under three built-in conservation scenarios: standard, contingency, and crisis. Users are also able to customize the tool to the specifics of their hospital and design custom conservation policies.

## Introduction

Substantial shortages of personal protective equipment (PPE) exist throughout the world due to the coronavirus disease 2019 (COVID-19) pandemic that has caused unprecedented surges in PPE demand. At the time of publication, several emergency measures have already been implemented to rapidly increase PPE supply and reduce the rate of its consumption.^3 4 7^ Yet, accurately forecasting how much PPE will be needed has been challenging, given the challenges in projecting how many patients will be hospitalized in different regions of the country.

During normal hospital operations, PPE procurement is a routine exercise of obtaining supplies at a fair price from reliable suppliers.^1^ Not much forecasting is needed – hospital administrators simply place replenishment orders for the materials they have consumed in the recent past. This creates a “pull” or “just-in-time” system in which the consumption of materials triggers orders for replacements.

Such pull systems may be optimal in normal conditions, as they allow hospitals facing significant cost pressures to be “lean” by operating with low inventory levels, but they do not support operations efficiently in exceptional situations such as a pandemic.^1^ Hospitals as well as local, state, and federal governments have created buffers in the form of “strategic stockpiles” that can help absorb unexpected spikes in demand resulting from such public health emergencies. However, forecasting the “right” PPE ordering quantities prior to the outbreak of a pandemic is practically impossible given the high variability in the number of people infected in different pandemics. The number of patients infected in past pandemics has ranged from thousands to millions during the SARS outbreak and the 1918 Flu pandemic, respectively.^2 6^

As a pandemic unfolds, more information about the pathogenicity of the virus as well as infection rates from other regions impacted earlier becomes available and can be used to inform epidemiological models forecasting infections, hospitalizations, and mortality rates. In the case of COVID-19, such epidemiological models include, among others, the COVID-19 Hospital Impact Model for Epidemics (CHIME) model from Penn Medicine^i^, the Institute for Health Metrics and Evaluation (IHME) Model from the University of Washington^ii^, and a model developed by a team at the University of Basel, Switzerland^iii^.

Though such hospitalization forecasting models are valuable for planning the need for beds and ventilators, they do not directly determine the demand for PPE. Limited data, variation in clinician behavior, and reduced, heterogeneous consumption in anticipation of PPE shortages make it challenging to translate patient volumes into forecasts of PPE demand. In this case study, we discuss the development and application of a tool for forecasting consumption of a set of PPE critical for the care of COVID-19 patients based on predicted patient admission and hospital census information. Our tool is publicly available to download at https://penn-chime.phl.io/. Our tool allows hospitals and health systems to make projections using three pre-populated scenarios. These scenarios—standard, contingency, and crisis— correspond to projections for PPE use by bedside clinicians (physicians, nurses, respiratory therapists) under increasingly strict PPE conservation policies. These scenarios were developed in consultation with clinicians across several different clinical departments and system-level leaders to ensure they capture realistic assumptions about how PPE materials are used in standard care within a hospital and what would constitute reasonable PPE conservation strategies in cases of PPE shortages. Our tool also provides users with the option to input their own custom scenarios, tailored to specific situations relevant to their hospital or health system. With the help of our forecasts, hospitals and health systems can update their PPE procurement decisions and adjust their consumption behavior by switching between the three scenarios or developing their own.

## Translating Patient Projections into PPE Demand Projections

Our forecasting tool requires daily forecasts for the expected number of COVID-19 patients who are hospitalized, the number in the ICU, and the number of new admissions. We then model the number of daily patients arriving in the ED as a multiple of daily new admissions. Our tool is flexible with respect to the source of these forecasts as long as their numerical values can be entered into an Excel Spreadsheet. The previously mentioned epidemiological models all allow for exporting this information and we are in ongoing discussions of directly integrating our tool with these models. A comparison of the three previously mentioned epidemiological models, including their methodology, key assumptions, and early predictive accuracy, can be found in Cotner et al.^4^

As we translate patient census into PPE demand, we need to explicitly model the clinical drivers of PPE consumption. Towards that goal, we found it helpful to distinguish between two types of PPE consumption: contact-based consumption and staffing based consumption. Contact-based consumption refers to PPE usage in which the number of items utilized is a direct function of the number of contacts staff members have with COVID-19 patients.

In contrast, staff-based consumption refers to PPE usage in which the number of items consumed only depends on the number of staff in a shift and not on the number of their contacts with patients. For example, masks in short supply are often now being used by a clinician for a shift, several days, or longer irrespective of the number of patient contacts unless the mask becomes visibly soiled or deformed. The number of masks needed therefore does not depend on the number of patient contacts but only on the number of staff on a shift. In contrast, clinicians typically change gloves when moving from one patient to another and a safe reuse of gloves is practically infeasible. Since every contact with a patient requires a new pair of gloves, we model them as contact-based consumption.

### Modeling Contact-based Consumption

Depending on the PPE item and the scenario, an item might be used only once (this practice is typical of the “standard” scenario) or might be reused multiple times (especially in scenarios of PPE shortage, such as “contingency” or “crisis”) before being discarded. The projected number of a particular PPE item (e.g., a pair of gloves) needed for a particular day (e.g., April 18) can be computed as:

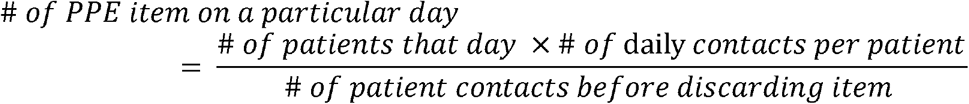

Our mathematical appendix provides a concise mathematical summary of all calculations. To illustrate, consider the number of pairs of gloves needed per day in an ICU with a census of 25 patients, assuming each patient is contacted by one clinician every hour, gloves are used at every contact, and gloves cannot be reused:

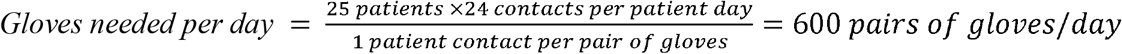

This formula does not require that the PPE be used for every patient contact. For example, if gloves were only used every *other* contact on average, then the denominator would be two patient contacts per pair of gloves because, on average, the patient would be contacted twice before a single pair of gloves is discarded. The total PPE demand for the hospital for a given item on a given day can be found by summing the demand across all portions of the hospital.

While these calculations by themselves are simple, obtaining the necessary data is not. In order to produce realistic values for the number of contacts with a patient, we relied on two different sources of information. One source was based on records of the number of times a nurse enters a patient’s room per day which we could calculate using badge tracking data. From this, we were able to extract the typical number of patient contacts per day for registered nurses (RNs) in some units. The second source of information was interviews with clinicians in the ICUs, regular medical floors, and Emergency Department to solicit information on how many times per day a typical patient is contacted by clinical personnel (RNs, residents, attendings, and respiratory therapists (RTs)).

The information we compiled from these sources was used to populate the assumptions in creating a “standard scenario.” To create values for the contingency and crisis scenarios, we asked clinicians to estimate the degree to which patient contacts could be reduced if PPE utilization needed to be mildly or sharply constrained (Table 1). These values do not represent a direct average across data sources, but rather a carefully considered weighing of different inputs. In order to produce realistic values for how PPE can be conserved through re-use (i.e., how many patients can be contacted using the same PPE item), we also surveyed clinicians, asking how often a piece of equipment is typically used under normal circumstances (standard scenario) and how many re-uses would be reasonable if resources were to become somewhat constrained (contingency) or extremely constrained (crisis). For equipment that is not used at every contact (e.g. *either* a surgical mask *or* a N95 is used) we relied on historical use data to find the relative frequency of use of such substitute items. The number of contacts before disposing of each equipment type under the different scenarios are shown in Table 2. Although we elicited values for all scenarios for all equipment, the staffing-based approach will be more appropriate for select equipment types (e.g., N95 masks).

**Table 1.**
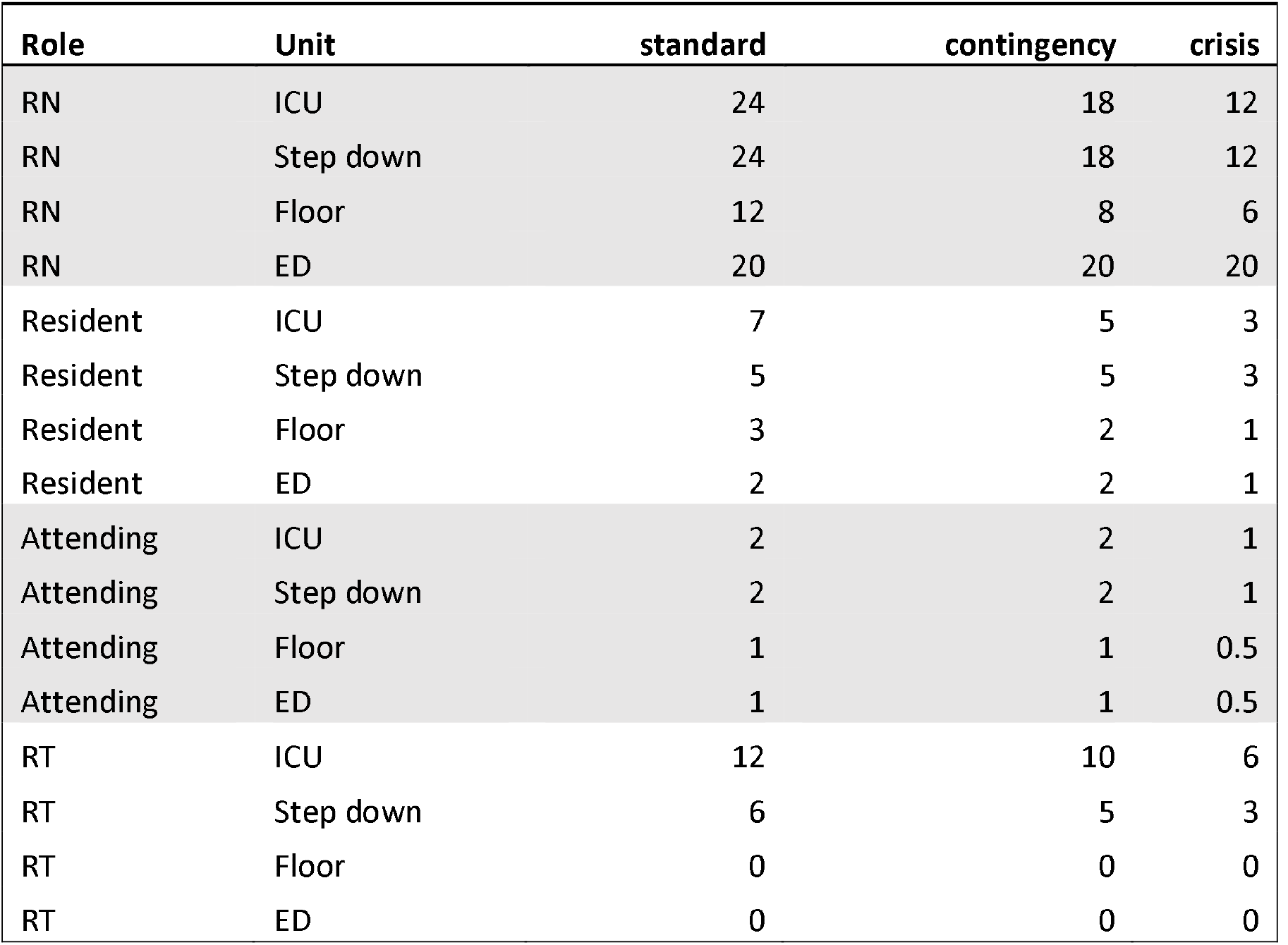
Assumptions about number of contacts per patient day for each scenario in our model. Users may also input custom values as needed.

| Role | Unit | standard | contingency | crisis |
| --- | --- | --- | --- | --- |
| RN | ICU | 24 | 18 | 12 |
| RN | Step down | 24 | 18 | 12 |
| RN | Floor | 12 | 8 | 6 |
| RN | ED | 20 | 20 | 20 |
| Resident | ICU | 7 | 5 | 3 |
| Resident | Step down | 5 | 5 | 3 |
| Resident | Floor | 3 | 2 | 1 |
| Resident | ED | 2 | 2 | 1 |
| Attending | ICU | 2 | 2 | 1 |
| Attending | Step down | 2 | 2 | 1 |
| Attending | Floor | 1 | 1 | 0.5 |
| Attending | ED | 1 | 1 | 0.5 |
| RT | ICU | 12 | 10 | 6 |
| RT | Step down | 6 | 5 | 3 |
| RT | Floor | 0 | 0 | 0 |
| RT | ED | 0 | 0 | 0 |

**Table 2.** Default parameter settings for the average number of contacts before discarding each type of equipment. While users always have the option to specify a contact-based policy for all equipment types, for some equipment types (e.g. gloves), our default scenarios assume staffing-based calculations.

| PPE | standard | contingency | crisis |
| --- | --- | --- | --- |
| N95 | 19 | 190 | 380 |
| Surgical mask | 1.05 | 10.5 | 20.1 |
| Glove | 1 | 3 | 5 |
| Gown | 1 | 3 | 5 |
| Disposable eye protection | 1 | 5 | 10 |

### Modeling Staff-based Consumption

In the case of staff-based consumption, rather than specifying that the clinician discard PPE after a given number of patient contacts, we instead specify that the equipment be (re)used for a certain number of shifts. For PPE items that are consumed based on staffing (as opposed to based on contacts), we can project the number of items of a particular PPE (e.g., a N95 mask) for a particular day (e.g., April 20) as:

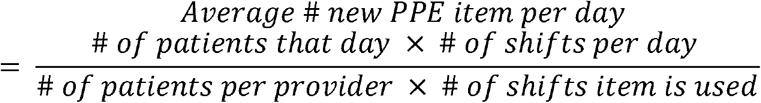

Staff-based calculations must be done separately for the multiple clinical roles (nurse, resident, RT), since staffing ratios vary across roles. To illustrate, consider the number of N95 masks needed per day in an ICU for nurses with a census of 20 patients. Assuming a patient-to-nurse staffing ratio of 2:1 and clinicians using a mask for five shifts before it is discarded:

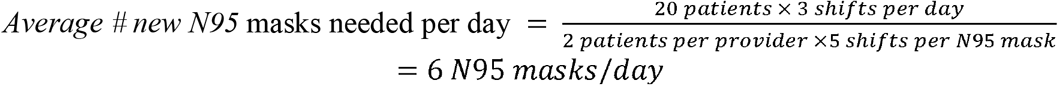

This calculation assumes that clinicians will be re-using the same mask from previous shifts. The total PPE demand for the hospital can be found by summing the demand across all clinical units and clinical roles. Tables 3–5 summarizes the values we used for each scenario in our calculations. Users also have the option to input custom values for each variable should the defaults not apply to their setting.

**Table 3.**
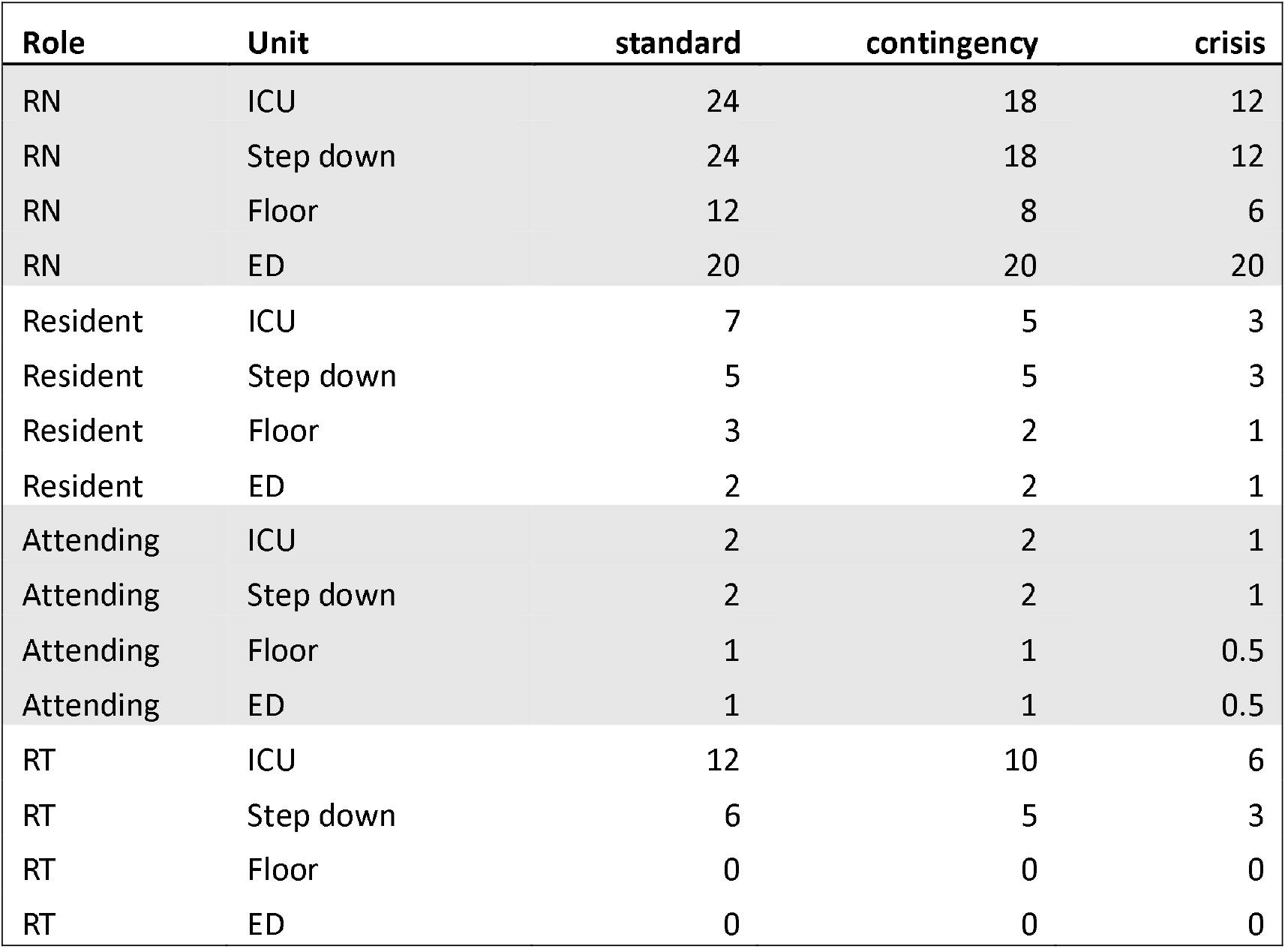
Default assumptions for patient to staff ratios. As always, users may input custom values.

**Table 4.** Default assumptions for number of shifts of use for each equipment type considered. As with the contact-based calculations, despite this table being populated for all equipment types and all scenarios, for some equipment types, we expect that a contact-based calculation will be more appropriate.

| PPE | standard | contingency | crisis |
| --- | --- | --- | --- |
| N95 | 1 | 10 | 20 |
| Surgical mask | 1 | 10 | 20 |
| glove | 1 | 3 | 5 |
| gown | 1 | 3 | 5 |
| Disposable eye protection | 1 | 5 | 10 |

**Table 5.** Default assumptions for shift lengths. We currently assume shift lengths are uniform across units.

| Shift length | hours |
| --- | --- |
| RN | 8 |
| Resident | 12 |
| Attending | 12 |
| RT | 8 |

As can be seen in the table, the values for staffing ratios, the levels of reuse, and even shift durations vary across the three scenarios. All values were obtained through discussions with system-level decision-makers as well as clinicians, where feasibility of re-use, patient and clinician safety, and equipment availability were considered.

### Scenario-dependent calculation bases

The consumption of some PPE items might be best modeled as contact-based in the standard scenario but as staff-based in cases of shortages (i.e., the contingency or the crisis scenario). For example, in our surveys, N95 masks in typical times are consumed based on the number of patient contacts. However, in light of supply shortages, many hospitals have switched utilization policies to a contingency mode limiting each staff member to one mask per shift, thus corresponding to a staff-based consumption. Table 6 describes how items are consumed either based on contact or based on staff across the three scenarios.

**Table 6.** Calculation type for each clinician and equipment type across units

|  | PPE type | standard | contingency | crisis |
| --- | --- | --- | --- | --- |
| ICU | N95 | contact | staff | staff |
| ICU | Surgical mask | contact | contact | contact |
| ICU | glove | contact | contact | contact |
| ICU | gown | contact | contact | contact |
| ICU | Disposable eye protection | contact | staff | staff |
| Step down | N95 | contact | staff | staff |
| Step down | Surgical mask | contact | contact | staff |
| Step down | glove | contact | contact | contact |
| Step down | gown | contact | contact | contact |
| Step down | Disposable eye protection | contact | staff | staff |
| Floor | N95 | contact | staff | staff |
| Floor | Surgical mask | contact | contact | staff |
| Floor | glove | contact | contact | contact |
| Floor | gown | contact | contact | contact |
| Floor | Disposable eye protection | contact | staff | staff |
| ED | N95 | contact | staff | staff |
| ED | Surgical mask | contact | contact | staff |
| ED | glove | contact | contact | contact |
| ED | gown | contact | contact | contact |
| ED | Disposable eye protection | contact | staff | staff |

## Model usage

We see multiple usages for this type of model. The most important during the pandemic would be for hospitals to assess needs for currently admitted patients and tie projections of hospitalized, ICU, and COVID-19 patients directly to the projected amount of PPE required to care for them. These predictions can be done using ‘standard usage’ assumptions.

A large number of COVID patients under standard assumptions would require a significant amount of PPE. For example, imagine a hospital will have 100 COVID ICU patients per day over ten weeks. During that time, for an item that requires contact-based use such as a pair of gloves, the hospital would need approximately 100 × 70 days x 24 contacts/day = 168,000 pairs of gloves just for the COVID ICU patients alone.

Estimates using standard assumptions can be adjusted based on existing or available inventory to contingency or crisis scenarios. If obtaining 168,000 gloves for the ICU (plus more for the regular floors, ED, and outpatient settings for COVID and non-COVID patients) seems infeasible, hospitals can estimate how many they would need under alternative scenarios so that utilization behavior can be adjusted before the shortage is critical, given that additional supplies may not be available at a later time.

As hospitals (and cities and states) may need to navigate through multiple waves of this epidemic, this tool can also be used to project future use for strategic stockpiling before potential future waves of the epidemic.

## Limitations

The biggest uncertainty of these projections comes from the epidemiological models that we use as our input. As we show in Cotner et al.^4^, these patient predictions vary widely and thus introduce noise into our PPE forecasts. Projections are more accurate within a 2–3 week horizon than further into the future, given the inherent uncertainty of predicting the exact course of the epidemic in different geographic regions.

We collected data and triangulated our estimates on actual PPE utilization through discussions with many clinicians within a single institution (Hospital of the University of Pennsylvania, Philadelphia, PA). Actual utilization patterns may differ at other hospitals or among other clinicians. Nevertheless, our model is flexible and allows for custom inputs based on utilization patterns at other facilities.

Norms about what constitutes “standard” utilization have been shifting due to widespread PPE shortages. Before the COVID-19 outbreak, it was standard to use a N95 mask in a wide range of clinical encounters. Due to shortages of N95 masks, their use in many health care settings is restricted to procedures that are likely to generate aerosols such as intubation, with surgical masks being recommended as the alternative for regular patient contact. Our assumptions on N95 versus surgical mask utilization were based on CDC recommendations and institutional policies at the University of Pennsylvania Health System. In addition, assumptions on reuse of N95 and surgical masks were based on surveys with clinicians on the feasibility of re-using each mask type (i.e., N95s are more durable than surgical masks) and the potential adoption of decontamination procedures (e.g., UV radiation) to prolong the mask’s useful product life.

Currently our model accounts for utilization in inpatient settings but does not include outpatient care or home health. Our model currently does not account for PPE use by non-clinical staff, such as cleaning staff. We also do not model needs for PPE in non-COVID patients since the PPE-use assumptions are focused exclusively on PPE-use for COVID patients. Health care systems using this tool should be careful to make sure that they account for regular utilization for non-COVID patients, for outpatient, and for other non-clinical roles that require PPE.

## Discussion

“Buffer or suffer” is an old saying among purchasing managers. Facing uncertainty, decision makers either have to be willing to hold what often appears to be unnecessary inventory or suffer severe shortages in cases of exceptionally high demand. In the case of PPE procurement, the uncertainty results from the unknown number of patients in the hospital as well as the heterogeneous consumption behavior of the staff in the hospital. Epidemiological models can help reduce the former uncertainty by modeling the arrival, admission, and census of patients in the hospital. The novelty of our approach is that it estimates the consumption of PPE as a function of the number of hospitalized COVID patients.

Though we developed our tool in the midst of the first wave of the COVID-19 pandemic, we believe this will have utility throughout the course of this pandemic. In addition, our model can be used in preparation for future public health emergencies and informing the strategic stockpiling decisions made by government officials. It can help identify expected shortages of particular PPE items that then can be mitigated by either increasing inventory reserves, pre-negotiating delivery agreements with PPE vendors, or changing utilization patterns among clinicians well in advance of critical shortages. Another use of our tool is to gain a better understanding of likely PPE consumption in the hospital over time. Though the procurement costs for PPE are not a major cost driver for clinical operations, they can easily add up to millions of dollars for a large hospital.

In normal times, procurement of PPE is fairly routine and can rely on ‘just in time’ delivery. During a pandemic planning ahead and using projections to estimate future needs is critical in mitigating risk to providers and patients by always having adequate stores of PPE on hand.

## Data Availability

All data used in this manuscript is given in the appendix.

## Mathematical appendix

### Contact-based calculations

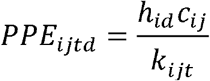

*PPE_ijtd_*: the projected number of PPE of type t (e.g. pairs of gloves) that will be used on day *d* by clinicians of type *j* (e.g., RNs) in unit *i* (e.g., the ICU)

*h_id_*: projected number of patients in unit *i* on day *d*

*c_ij_*: the average number of times each patient is contacted in a 24-hour period by a clinician of type *j* in unit *i*

*k_ijt_*: average number of contacts before *t* is discarded by clinicians of type *j* in unit *i*. For disposable items that are used at every contact under standard conditions, this value is one. For disposable items that are on average used every nth contact (and discarded after each use) this value is *n*; if an item is re-used *n* times, the value is also *n*.

### Staffing-based calculations

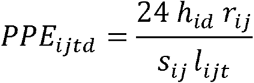

*PPE_ijtd_*: the projected number of PPE of type t (e.g. pairs of gloves) that will be used on day *d* by clinicians of type *j* (e.g., RNs) in unit *i* (e.g., the ICU)

*h_id_*: projected number of patients in unit *i* on day *d*

*r_ij_*: the patient to clinician ratio for clinicians of type *j* on unit *i*

*s_ij,_*: shift length in hours

*l_ijt_*: the number of shifts for which clinicians of type *j* in unit *i* will use PPE of type *t* before discarding

## Footnotes

i https://penn-chime.phl.io/

ii https://covid19.healthdata.org/united-states-of-america

iii https://covid19-scenarios.org/

